# Analysis, quantification, and visualization of RT-LAMP technique for detection of COVID-19

**DOI:** 10.1101/2021.07.15.21260528

**Authors:** Mohammad H Ghazimoradi, Maryam Daryani, Masoud Garshasbi, Ehsan Zolghadr, Ali Khalafizadeh, Sadegh Babashah

## Abstract

**Background:** SARS-Cov-2 is a new virus that caused an epidemic disease, COVID-19. According to the world health organization, detecting the patients/carriers is by the far the most important action to prevent the pandemic. Recently, the loop-mediated isothermal amplification (LAMP) technique has become more popular due to the easy handling of a one-step kit used for the detection of many diseases than RT-PCR-based techniques.

**methods:** Herein, we used the RT-LAMP technique so as to detect COVID-19. To this end, 40 paired-samples of patients and healthy people had been collected and tested by RT-PCR for *N* and *E* genes of SARS-CoV-2. The RT-LAMP test has been performed on samples for the *RdRp* gene. The sensitivity and specificity of tests have been determined.

**Results:** The testing results are consistent with the conventional RT-qPCR. Additionally, we also showed that a one-step process without RNA extraction is feasible to achieve RNA amplification directly from a sample.

**Conclusion:** We confirmed that RT-LAMP is a rapid, simple, and sensitive method that can be used as a large-screening method, particularly in regional hospitals with limited access to high-technologies.

## Introduction

Coronaviridae contains different obnoxious viruses that have caused many epidemics, e.g. respiratory syndrome (SARS-CoV) and more recently the Middle East respiratory syndrome (MERS-CoV) ^1,2^. Unfortunately, crossing the species barrier of this family caused some pandemics. On top of that, as a new disease, acute respiratory syndrome coronavirus 2 caused COVID-19 that tremendously affects the human population ^3^.

As conventional wisdom, by far the most effective way to prevent COVID-19 is social distancing and quarantine which would stop its spreading within populations ^4^. To this end, detecting infected individuals is a part-and-parcel process. Generally, RNA viruses can effectively be isolated and detected by real-time (RT)-PCR ^5,6^. Unfortunately, this method is somehow cost-incurring requiring expensive equipment that is not easily accessible in many parts of the world ^7^. Besides, the capacity of this technique is limited to many samples ^8^. Hence, there is an urgent need to introduce a high-throughput technique addressing such issues.

In the past decade, various isothermal amplification techniques were established that are contain recombinase polymerase amplification, rolling circle amplification, and helicase-dependent amplification. Revolutionary, Notomi *et al*. introduced a new isothermal amplification method that was capable of amplifying a limited amount of DNA copies in a short time, i.e. in an hour ^9^. This method is called “*loop-mediated isothermal amplification (LAMP).”* In contrast to PCR-based techniques, new isothermal techniques—e.g. Loop-Mediated Isothermal Amplification (LAMP)—do not need the prolonged temperature cycles ^9,10^. LAMP uses a different set of primers, e.g. a set of four (or six) different primers binds to six (or eight) different regions on the target gene making it highly specific, so as to amplify DNA fragments without using any complicated instruments ^10,11^.

Like RT-PCR, RT-LAMP uses reverse transcriptase to make complementary DNA from RNA templates and is further amplified by using DNA polymerase ^12,13^. This method is beneficial because the assay can be completed in a single step, by incubating all the primers and enzymes (Bst polymerase and reverse transcriptase) with a constant temperature ^14^, thus this method is effective in detecting RNA viruses in a pretty short time. According to Wang *et al*., in addition to helping in diagnosis, the RT-LAMP can also be used as an epidemiologic surveillance system for human virus infections, e.g influenza, Hepatitis C, Ebola, respiratory syncytial, zika virus, and MERS-CoV ^15^. Currently, one of the most successful uses of RT-LAMP is diagnosing the HIV retrovirus that showed the sensitivity of 670 viruses per µl of whole blood ^16^. Damhorst *et al*. suggested RT-LAMP as a function for controlling hospital-based infections and for laboratory-based observations ^16^. As a consequence, visual inspection alleviates the need for gel electrophoresis, reduces the assay time, and thus, makes the method suitable for field tests ^17,18^.

LAMP PCR uses different sets of primers that involve two outer (F3 and B3) primers, two inner primers— forward (FIP) and backward inner primer (BIP)—and loop primers that per se can increase the specificity. By modifying LAMP, RT-LAMP has been extended to use a reverse transcriptase instead ^19^. Besides, *Bcabest* is an enzyme used in RT-LAMP that can synthesize cDNA from RNA directly ^20,21^. The present study aimed to determine the possibility of using the RT-LAMP method for detecting COVID-19 in 40 confirmed patients.

## Materials and methods

### Sample collection

Blood samples were taken from 40 confirmed infected patients with Sars-Cov-2 that had been previously verified using RT-qPCR at DeNA Laboratory, Tehran, Iran. We also collected 40 samples from whom their PCR test was negative for Sars-Cov-2 (healthy group). RT-qPCR was conducted to confirm each sample. The study received ethical clearance from the ethical committee of Tarbiat Modares University, Tehran, Iran. COVID-19-infected whole blood samples were specimens left over from the diagnostic laboratory and were unlinked from personal identifiers before use (CDC IRB protocol #1896).

### RNA extraction

Total RNA was extracted from each sample using the QIAamp Viral mini kit (Qiagen, Il, USA) according to the manufacturer’s instructions. In order to investigate and exclude this notion that the primers may cause unspecific products using human RNA as the template and determine the sensitivity of the RT-LAMP and PCR reaction for DNA, we extracted RNA samples of *human colorectal cells* derived from the patients with colorectal cancer using TRIzol (Sigma, TX, USA). To be on the safe side, we measured the concentration and purity ratio of the isolated RNA samples using a Nanodrop™ 2000c (Thermo Fisher Scientific, IL, USA). The samples were also run at 1% agarose gel for further validation. In order to cut down on any possible DNA contamination, the samples were treated with RNase-free DNase (Qiagen, Valencia, CA). In brief, the test contained three probe-base primers for *N* and *E* genes of Sars-Cov-2. Another probe was used to determine the human *RNase P* gene as the control.

### RT-qPCR

In order to reevaluated and reconfirm all patients’ status using RT-PCR, isolated RNAs were reverse-transcribed using the miR-Amp Kit (ParsGenome co., Tehran, Iran) and RT-PCR was conducted according to the standard protocols ^22,23^. In brief, RT-qPCR amplification of the *SARS-CoV-2 N protein* gene was performed with the following sets of primers: the N-1 set, the forward primer 5′-CACATTGGCACCCGCAATC-3′, the reverse primer 5′-GAGGAACGAGAAGAGGCTTG-3′, and the probe 5′-FAM-ACTTCCTCAAGGAACAACATTGCCA-TAMRA-3′ (where FAM is 6-carboxyfluorescein and TAMRA is 6-carboxytetramethylrhodamine), and the N-2 set, the forward primer 5′-AAA TTT TGG GGA CCA GGA AC-3′, the reverse primer 5′-TGGCAGCTGTGTAGGTCAAC-3′, and the probe 5′-FAM-ATGTCGCGCATTGGCATGGA-TAMRA-3′

### Primer design and RT-LAMP reaction

Primers were designed using Primer3.0 online tool ^24^ against the *RdRp* gene in ORF-1b of Sars-Cov-2. The sequence of primers was as follow: F3: 5’-TGTTTTATTGCCACTAGTCTCT-3’, B3: 5’-ACATGTATAGCATGGAACCAA-3’, FIB: 5’-ACGTGTGAAAGAATTAGTGTATGCAGTCAGTGTGTTAATCTTACAACC-3’, BIB: 5’-CCTGACAAAGTTTTCAGATCCTCAGTAACATTGGAAAAGAAAGGTAAG-3’. The BecaBest enzyme was used and the PCR was conducted according to the following conditions: 1 min at 65°C, 5 min at 30°C, 30 min at 65°C, 5 min at 98°C as the final extension step, and 1 min at 5°C. The PCR products were run on a 2% agarose gel at 90 volts for 90 minutes. Negative controls were included in each run, including a water control to check for cross-contamination. After gel electrophoresis, the visualization of results was done under Ultra Violet (UV) light on a 1.2% agarose gel, followed by staining with ethidium bromide.

### Qubit, gel electrophoresis, and visualization

Qubit has been performing by dsDNA Hs assay kit (Thermofisher, TX, USA). In this method, the double-strand DNA is detected by fluorescence molecules that only attach to the double-strand DNA. Gel electrophoresis has been performed using 2% agarose gel. For visualization, Kbc Power Load Dye (CinnaGen Co., Iran) was used.

### RT-LAMP specificity

Amplification specificity was determined by sequencing the products using Sanger sequencing. To this end, the LAMP products were extracted from agarose gel using the QIAquick Gel Extraction Kit (Qiagen, Germany). Sequencing was performed with ABI 3730 DNA Sequencer (Applied Biosystems, Foster City, CA, United States) by capillary electrophoresis. Sanger sequencing results were analyzed using Chromas Lite v 2.01 (Technelysium Pty Ltd., Tewantin, QLD, Australia) to confirm the data.

## Results

### RT-qPCR

RNA samples were isolated from 40 Sars-Cov-2 infected patients. These samples were reconfirmed regarding the effective history with Sars-Cov-2 by RT-PCR (**figure 1A and B Supplement**). The samples were positive for both genes of *N* and *E* of Sars-Cov-2. The average of CT was determined 29.86±6.28 (**Figure 1A**). To validate amplification, PCR-products were analyzed using Qubit *vs*. non-amplified samples (**Figure 1B**). Afterward, samples were visualized by running the products via electrophoresis on agarose gel (**Figure 1C**).

**Figure 1.**
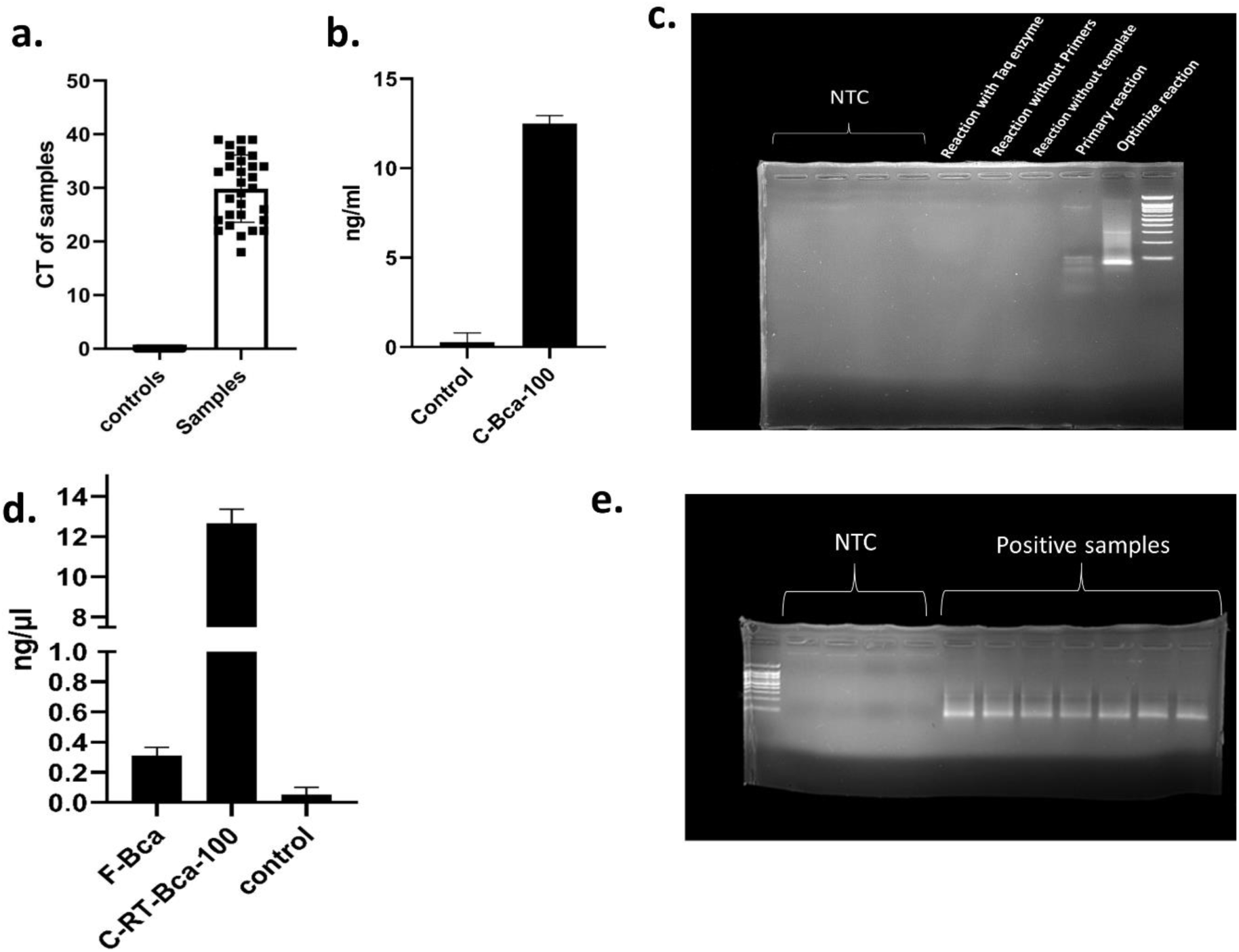
Set up of LAMP technique. **a)** CT of samples versus controls. **b)** Quantities of determination of reactions. **c)** Gel electrophoresis of the products (C-BCA-100= 1,100 ng of Coronavirus RNA amplified by Bcabest enzyme). **d)** Quantification of RNA and DNA polymerase activity of reactions (F-BCA= 100 ng of Coronavirus RNA amplified by Bcabest enzyme with only inner primer set, C-RT-BCA-100= 100 ng of Coronavirus RNA amplified by Bcabest enzyme in a complete solution for complete reverse transcriptase and amplification activity). **e)** Replication of samples and controls with 100 bp ladder.

### RT-LAMP

Since BcaBest enzyme is a polymerase that can also amplify DNA as well, to know the involvement of each activity, a reaction with F1B and F2b primer was performed to measure reverse transcriptase *vs*. polymerase activities (**Figure 1D**). For further confirmation, 40 samples have been replicated (**Figure 1E**) using RT-qPCR. Clear bonds have been observed after electrophoresis. For validation of this step, the DNA samples were extracted from the agarose gel using QIAquick Gel Extraction Kit (QIAGEN, TX, USA) and PCR was performed by B1 and F1 primers. The products were sequenced by the Sanger sequencing ABI 3730 instrument. In order to validate the products of LAMP reaction/sensitivity, bi-directional Sanger sequencing was performed (**Figure 2A and B**).

**Figure 2.**
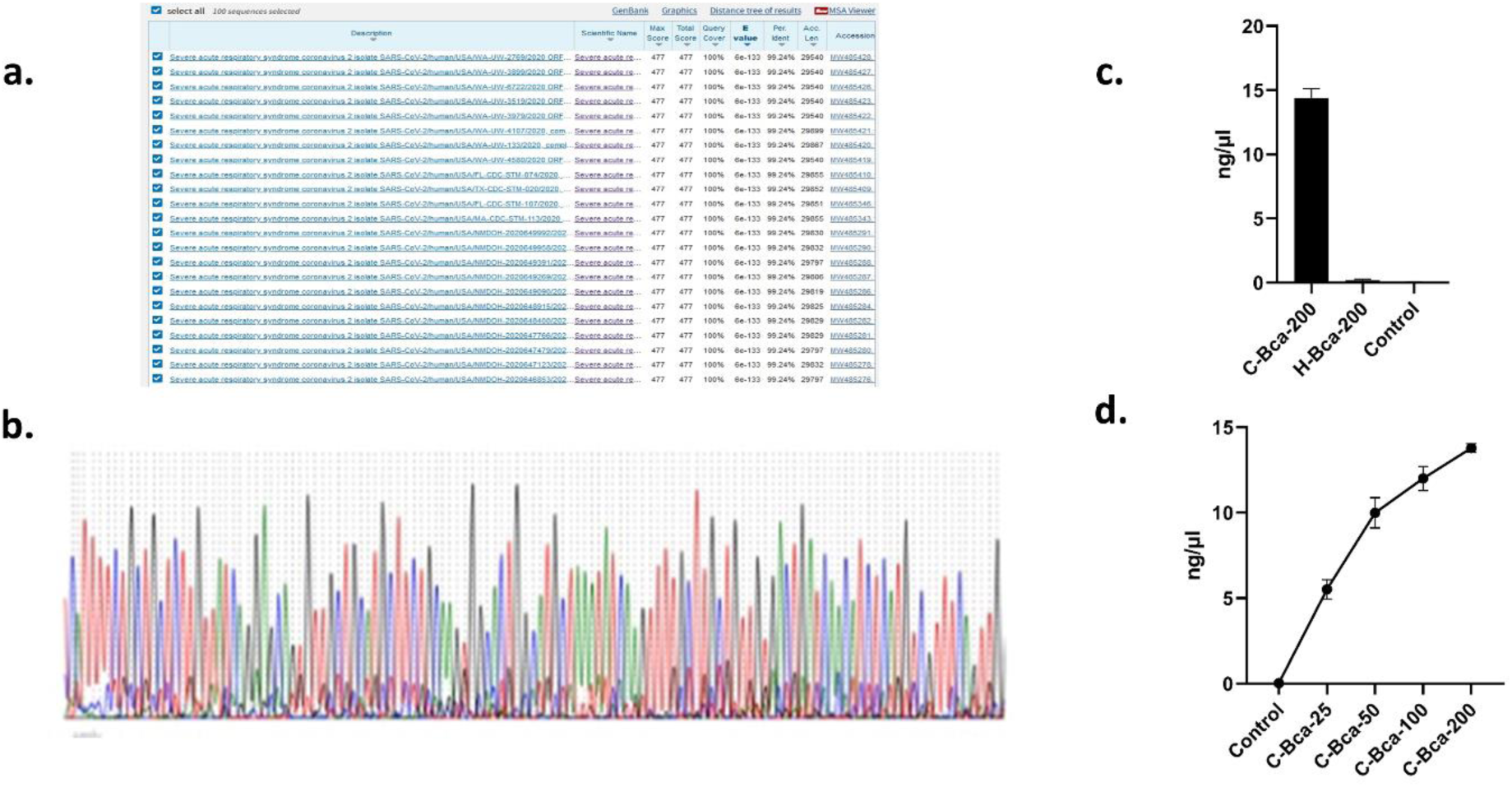
Validation, sensitivity, and specificity of the RT-LAMP for COVID-19. **a and b)** sequencing and the blast of the amplified fragment as validation of specific amplification. **c)** sensitivity test for determination of the fewest doses of the viral load (C-BCA-25, 50, 100, and 200= 25, 50, 100, and 200 ng of Coronavirus RNA amplified by Bcabest enzyme). **d)** specificity test for primer test (H-BCA-200= 200 ng of human RNA amplified by Bcabest enzyme, C-BCA-200= 200 ng of Coronavirus RNA amplified by Bcabest enzyme).

### Evaluation of the diagnostic performance of RT-LAMP

Different concentration of RNA was added to reaction so as to investigate the sensitivity; we observed that concentration more than 100 µg generate a flat phase (**Figure 2C**). Furthermore, adding human RNA to the reaction would not lead to any amplification (**Figure 2D**). Last but not least, all products were well-recognizable under UV light, suggesting that this technique is handy for COVID-19 detection (**Figure 3 A**). A diagnostic evaluation test was performed to compare RT-LAMP and RT-PCR as the gold standard technique for the diagnosis of COVID-19. According to the result, the sensitivity of the RT-LAMP was determined as equal as RT-qPCR that was performed for the diagnosis of COVID-19. In other words, neither false nor positive result was identified using RT=qPCR and RT-LAMP.

**Figure 3.**
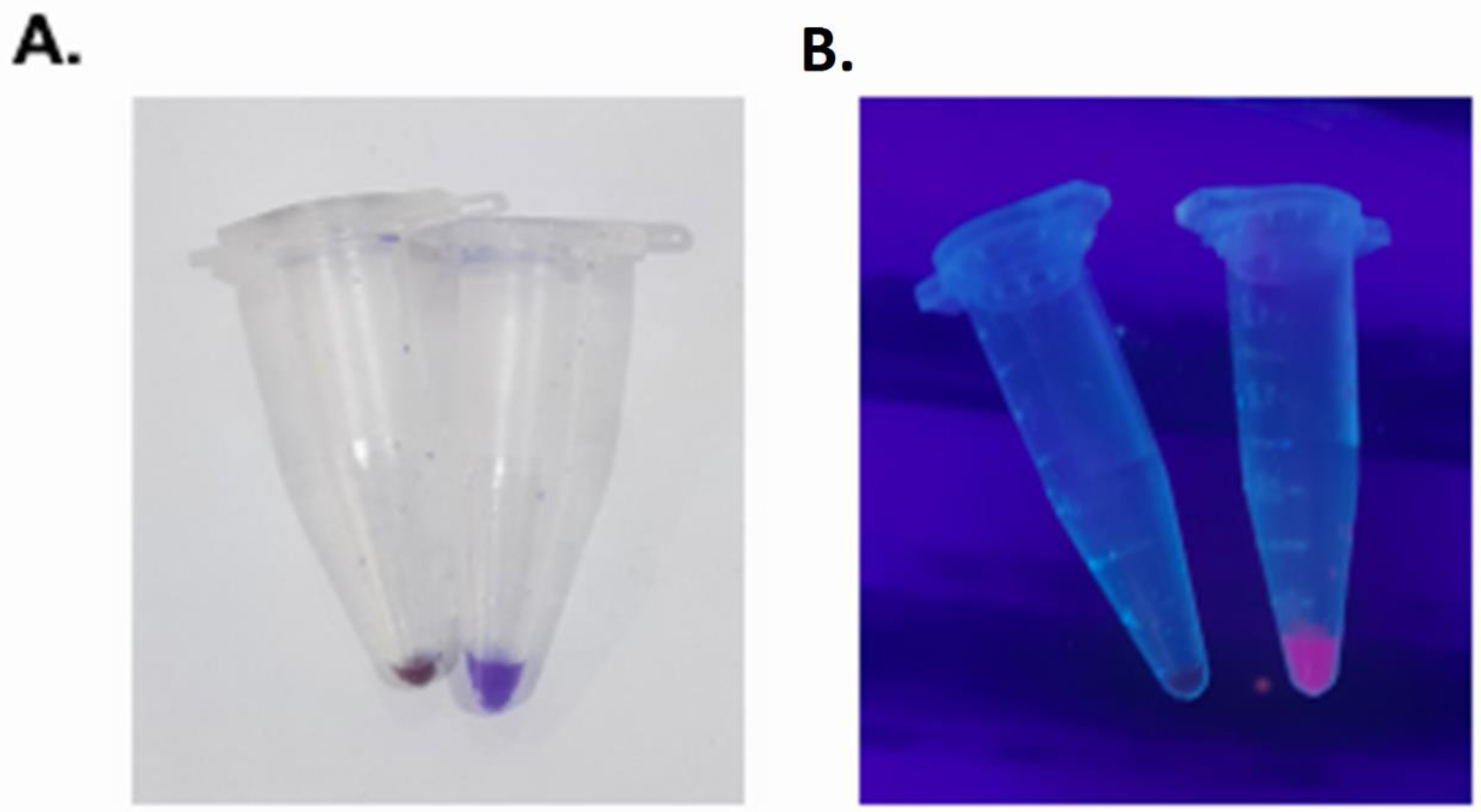
**a)** demonstration of reaction mix with power loading buffer for the naked eye. B) demonstration of reaction mix with power loading buffer under UV light.

## Discussion

A cost-effective diagnostic or confirmatory test is required for the detection of COVID-19 infection in a short time. Due to the false-negative result of daily used techniques ^25,26^, the spreading cannot be controlled very well. On top of that, the current techniques are needed some high degree of technical skill and time required for the assay ^26^. Therefore, in the present study, we aimed to determine the possibility of RT-LAMP application in COVID-19 detection.

LAMP technique is one of the most trusted techniques that are frequently used as a rapid test for detection of diseases and/or pathogens; this technique does not require any expensive instrument, above and beyond that, it is not a cost-incurring but with the high rate of specificity than traditional PCR-based techniques ^27^. Herein, we used the RT-LAMP technique to detect COVID-19 in patients; this technique showed a good amplification of around 10 to 15 ng/µl. The sensitivity and specificity of the LAMP test for COVID-19 detection were investigated by showing a high sensitivity to human RNA and good resistance to a low amount of target RNA. This technique could also successfully produce cDNA from viral RNA and amplify it. Using only F1B and F1c primers, show a 0.1 ng/ml which is lesser than 12 ng/ml of complete reaction.

The adaptation of the LAMP technology for the detection of COVID-19 infection has many advantages: first and foremost, LAMP occurs under isothermal conditions ^28^, thus it does not require a separate denaturing step; secondly, the needs for any special equipment, such as a thermal cycler, is alleviated and reactions can be accomplished in a regular heat block or water bath ^29^; besides, we used the six pair of primers that can, in turn, decrease the false-positive results that are frequently reported in other tests. Using the primers can also increase specificity *ipso facto*. Additionally, the amplified LAMP products can be visually detected by turbidity because of the accumulation of the magnesium-pyrophosphate by-product. As well, due to the large amplificated regions, they can be traced using DNA staining processes that help us to replicate the results as precise as possible. Some LAMP techniques have been recently generated ^30^, which can be used world-widely and easily. One of the important benefits of using RT-LAMP is that LAMP exhibits less sensitivity to inhibitory substances present in biological samples than PCR ^19^.

In this study, we adapted the RT-LAMP assay to detect the target sequences, i.e. *N* and *P* genes in Sars-Cov-2. Though the primers were designed against such specific genes, recognition of clinical isolates was confirmed using patient blood samples. All steps were also validated using Sanger sequencing as the gold standard of sequencing processes. The sensitivity of the RT-LAMP for COVID-19 RNA was determined to be in a range from 20 to 200 µg of the templates when using a 25 µl final reaction volume.

As the patients in the initial stages have a lower viral load, this can put the test on the brink of low-detection rate; to avoid, the sensitivity of the assay can be increased by increasing the overall reaction mix volume (>25 µl) as postulated by previous studies ^31^. Since there is an urgent need to detect Covid-19 as soon and accurately as possible, we introduced the RT-LAMP technique. This technique is an easy, one-pot, and rapid method that its products can be visualized by the naked eye. Besides, this technique shows significant specificity and sensitivity in comparison with conventional methods such as RT-PCR. In sum, this method can perfectly detect COVID-19 in low-income countries and countries with weak medical infrastructures.

## Data Availability

All of the data has been demonstrated and available in manuscript.

## Availibility of data

Not applicable.

## Code Availibility

Not applicable.

## Ethics approval

This study received ethical clearance from the ethical committee of Tarbiat Modares University, Tehran, Iran.

## Consent to participation

All of the authurs declare that this publication has been send for publication.

## Authur contribiotion

Mohammad H Ghazimoradi was responsible for conniving idea, experiment design and performing experiment. Maryam Daryani was responsible to clinical and statical evaluation of work. Masoude Gharshasbi participate in sample collection. Ehsan Zolghadr and Ali khalafizadeh were responsible for wright the manuscripts. Sadegh Babashah provide management and financial aid of work.

## Acknowledgment

We thank Dr. Fatemeh Bitarafan for her help in the preparation of this article.

---

**Figure 1S.**
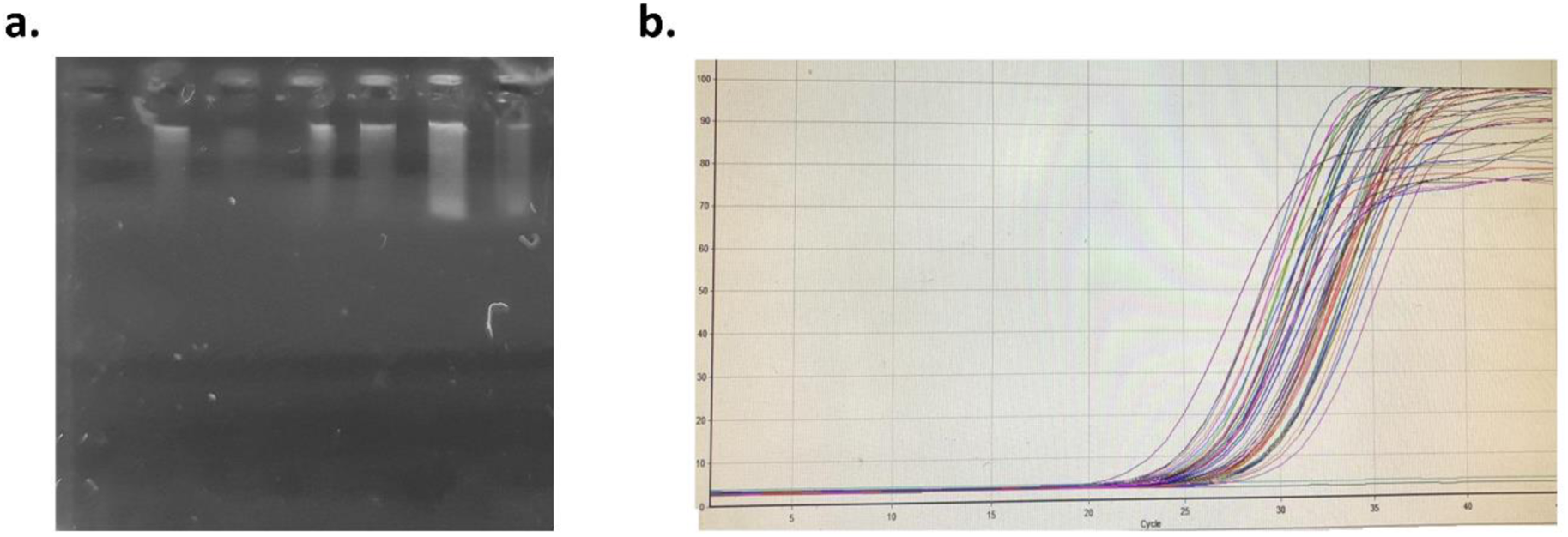
The validity of collected samples. **a)** Gel electrophoresis of extracted RNA from infected samples. **b)** RT PCR of samples as a validation of infection.

